# Skeletal muscle and intermuscular adipose tissue gene expression profiling identifies new biomarkers with prognostic significance for insulin resistance progression and intervention response

**DOI:** 10.1101/2022.01.07.21268359

**Authors:** Dominik Lutter, Stephan Sachs, Marc Walter, Anna Kerege, Leigh Perreault, Darcy E Kahn, Amare D Wolide, Maximilian Kleinert, Bryan C Bergman, Susanna Hofmann

## Abstract

Although insulin resistance often leads to Type 2 Diabetes Mellitus (T2D), its early stages remain often unrecognized thus reducing the probability of successful prevention and intervention. Moreover, treatment efficacy is affected by the genetics of the individual patient. We used gene expression profiles from a cross-sectional study to identify potential candidate genes for the prediction of diabetes risk and intervention response. Using a multivariate regression model, we linked gene expression profiles of human skeletal muscle and intermuscular adipose tissue (IMAT) to fasting glucose (FG) and glucose infusion rate (GIR). Predictive potential of identified candidate genes was validated with muscular gene expression data from a longitudinal intervention study. We found that genes with a strong association to clinical measures clustered into three distinct expression patterns. Their predictive values for insulin resistance varied strongly between muscle and IMAT. Moreover, we discovered that individual gene expression based classifications may differ from those classifications based predominantly on clinical parameters indicating a potential incomplete patient stratification. Out of the 15 top hit candidate genes, we identified *ST3GAL2, AASS, ARF1* and the transcription factor *SIN3A* as novel candidates for a refined diabetes risk and intervention response prediction. Our results confirm that disease progression and a successful intervention depend on individual genetics. We anticipate that our findings may lead to a better understanding and prediction of the individual diabetes risk and may help to develop individualized intervention strategies.

## Introduction

Obesity is a frequent precondition for the development of chronic metabolic diseases like insulin resistance (IR) and type 2 diabetes (T2D). Based on the recently published results from the 2017– 2018 National Health and Nutrition Examination Survey (NHANES) 42.5% of U.S. adults are currently obese and are thus all individuals at high risk for developing T2D and its complications [1]. Moreover, the International Diabetes Federation (IDF) predicts an 51% increase from 463 to 700 million individuals with diabetes worldwide by 2045 and indicates that 1 in 2 adults with diabetes remain undiagnosed at presence [2]. Although current assessment of diabetes and prediabetes is based on purely glycemic indicators it is important to emphasize that the risk for developing diabetes is also dependent on age, sex, fat tissue distribution, genetic, environmental, and ethnic characteristics. Depending on these individual risk factors and on the inclusion criteria of the studied cohorts a wide heterogeneity in the progression from prediabetes to diabetes has been observed. Emerging evidence from a population-based study with 381,363 participants indicates that even people referred to as metabolically healthy obese are at a substantially higher risk to develop diabetes and its complications [3]. Although interventions, medically or by change of lifestyle (diet, exercise) reduce the risk for severe complications evidence is emerging in population-based cohorts that treatment efficacy also depends on individual’s genetics [4-6]. Meaning that patients treated with glucose-lowering interventions vary in their response with some gaining a considerable benefit, others no benefit; and some getting limiting side effects. The prevalent opinion in the field is that there are many factors impacting this variation, but one key factor is a heritable trait [7]. Taken together, it is becoming increasingly clear that the current clinical standards to define the metabolic health status of an individual is not adequate and new strategies for the effective prevention of diabetes have become critically important to reduce the impact of this disease burden. A deeper understanding of the individual features and a precise phenotyping of prediabetes may improve stratification of disease risk and optimize the benefit/risk ratio and cost-effectiveness of any therapeutic approach for the prevention, intervention and improvement of T2D.

Given that skeletal muscle is responsible for more than 85% of insulin-stimulated whole-body glucose disposal [8], and that any dysfunction impairing glucose metabolism in this tissue will affect whole-body glucose homeostasis ultimately contributing to the development of diabetes [9] mechanistic studies mostly focus on this very tissue to elucidate metabolic adaptation and its regulation. More recently, evidence is pointing to intermuscular adipose tissue (IMAT) accumulation as another local regulator of muscular IR and the progression to diabetes [10, 11]. We thus hypothesized in this study that tissue specific gene expression profiling of muscular and/or IMAT can achieve a more specific and detailed characterization and classification of individual physiological states than circulating parameters alone. We further presumed that the expression of individual genes might (i) allow for the prediction of individual disease related states, (ii) identify individuals with a high or low risk for diabetes, and (iii) assess beforehand the potential response of a given individual to a specific treatment strategy.

To this end, we investigated herein interactions between gene expression and clinical diabetes markers in skeletal muscle and IMAT from overweight individuals with and without diagnosed T2D. We used multivariate regression to model the tissue specific gene expression impact on the two key IR markers, glucose infusion rate (GIR) during a hyperinsulinemic/euglycemic clamp and fasting glucose (FG). Out of the 65 top predictive genes, we identified three distinct gene clusters. Gene expression-based classification of our study participants led to a refined view on an individual’s metabolic state that partly differs from the clinical classification. In an independent lifestyle and exercise intervention study with diabetic patients, we could validate some selected candidates and identify four genes with the potential to predict individual intervention responses.

## Methods

### Human transcriptional profiling dataset

Human muscle and IMAT transcriptional profiles were taken from a cross-sectional study previously published by Sachs et al. [12]. To identify features suitable for characterization of individual pre-diabetic states and potentially predictive for disease progression we selected all 16 participants with obesity (OB) and diabetes mellitus type two (T2D) for which paired samples were available. All participants were clinically characterized by determining insulin sensitivity via glucose infusion rate (GIR, mg/kg/min) during hyperinsulinemic-euglycemic clamp, fasting glucose (FG, mg/dl), age, BMI, relative fat mass (RelFat, % kg), fat free mass (FFM, kg), height (in), body weight (BW, kg) (Table 1, Figure 1AB, S1AB).

**Table 1.**
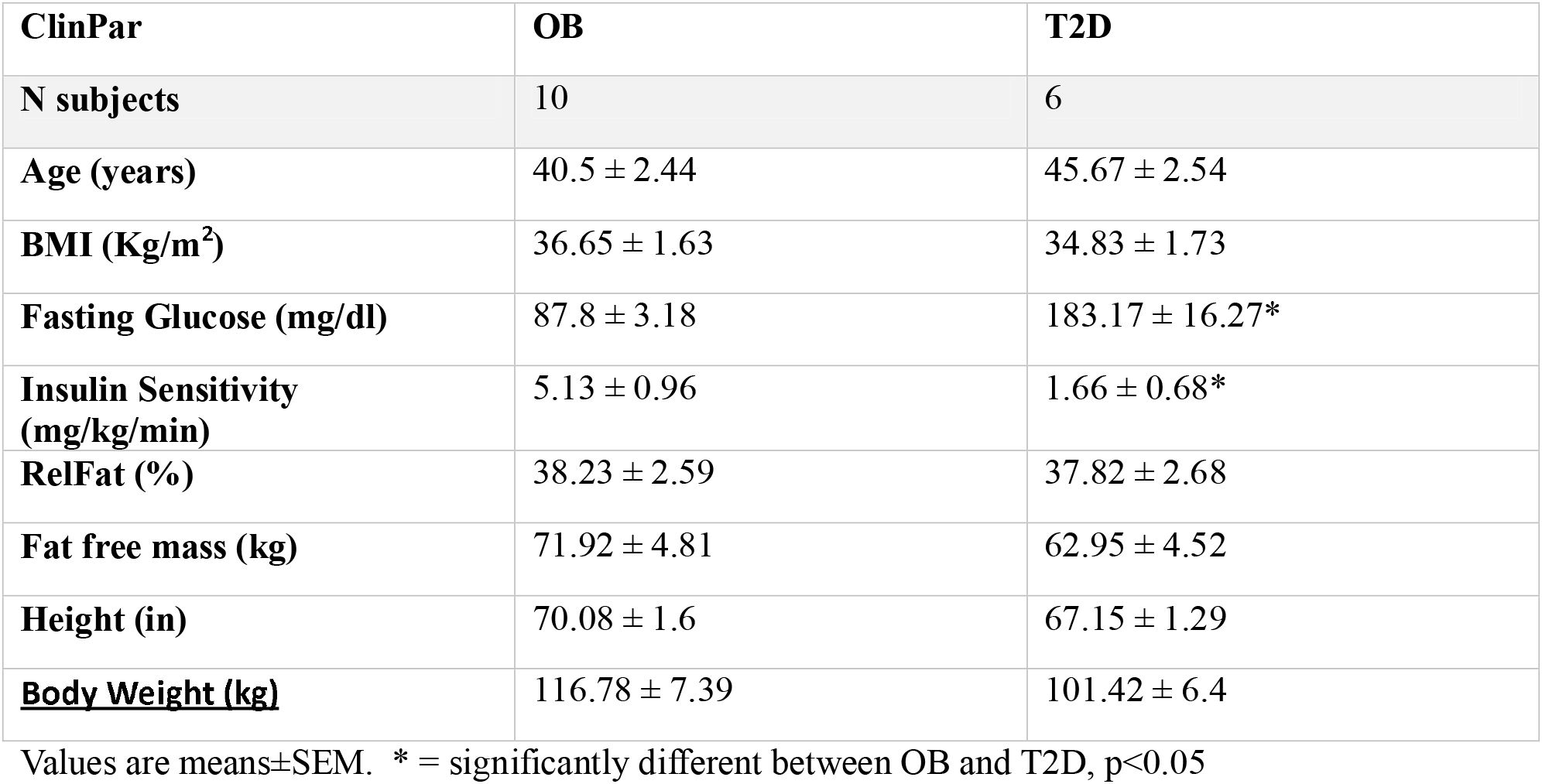
Subject demographics: Human transcriptional Profiling (N = 16)

**Figure 1:**
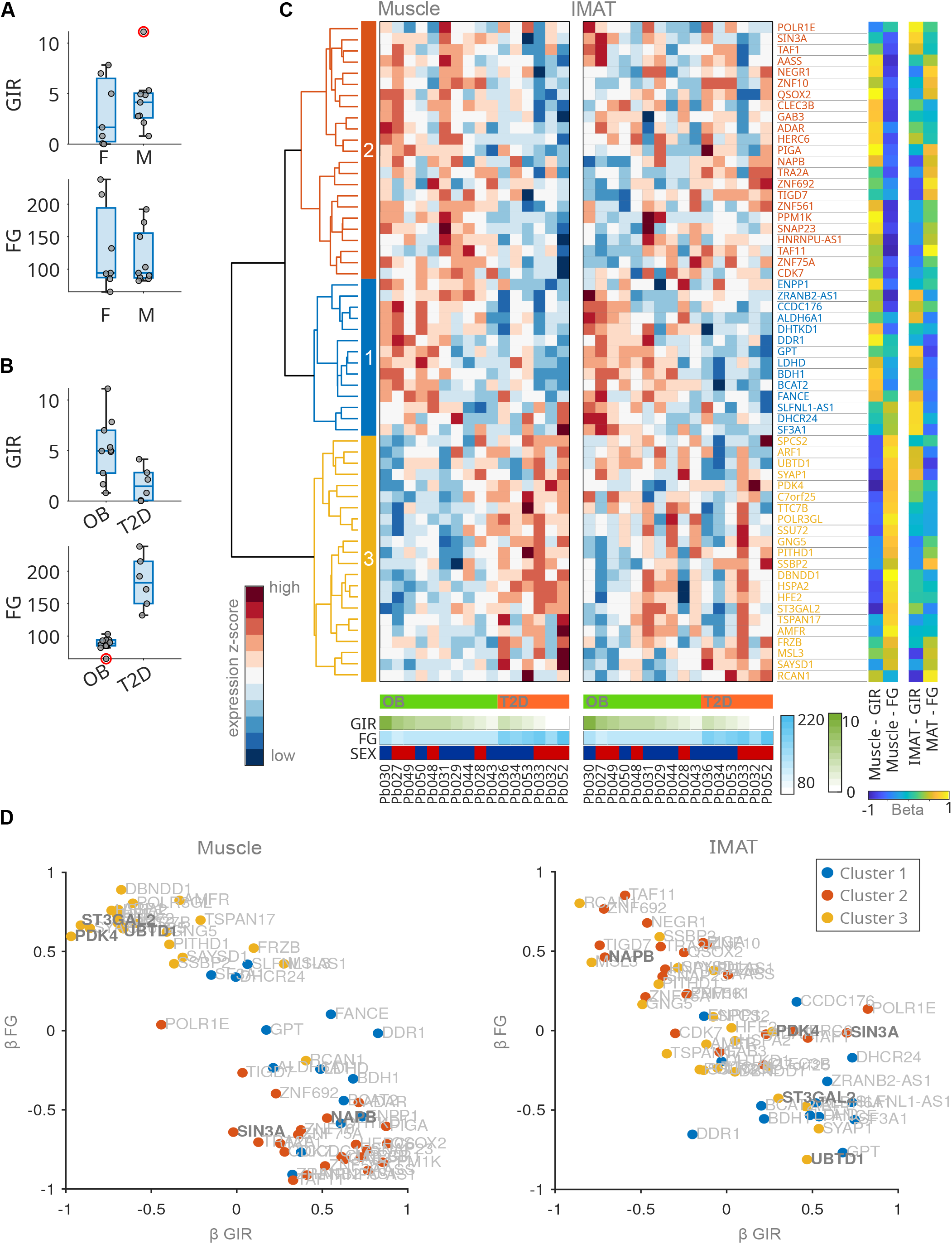
Multivariate regression analysis unravels tissue specific gene expression patterns correlating with insulin resistance. (**A, B**) Boxplot comparing Fasting Glucose and Insulin sensitivity (GIR mg/kg/min) distributions between subjects of different gender (A) and classifications (B). OB: obese, T2D: type 2 diabetic F: female, M: male. Red marked dots indicate outliers. (**C**) Heatmaps of muscle and IMAT genes correlating with insulin resistance identified by multivariate regression. Colors of left dendrogram refer to identified clusters 1-3. Bottom color bars indicate individual disease classification, Glucose Infusion Rate (GIR), Fasting Glucose levels and sex. Vertical color bars show the four estimated regression coefficients for each gene indicating four relationships between tissues and response variables (GIR & FGl). (**D**) Scatterplots comparing gene specific regression coefficients β for GIR and FGl for muscle and IMAT tissue. Dot colors refers to gene cluster assignment.

### Longitudinal Intervention dataset

Seventeen individuals with obesity with and without pre-diabetes were recruited for this study. Subjects were asked to refrain from planned physical activity for 48 hours before the metabolic study and were given a standardized diet for 7 days prior to the first and second metabolic study. After overnight fasting a basal muscle biopsy was taken and followed by metabolic profiling including a 3-hour hyperinsulinemic euglycemic clamp. After the first metabolic study, volunteers entered a 12-week supervised weight loss and exercise training intervention. After the 3-month intervention, subjects transitioned to a 2-week weight maintenance diet. Volunteers continued to exercise during the weight stabilization period. After completing the intervention and 2-week weight maintenance period volunteers then repeated the metabolic study with the muscle biopsy and 3-hour hyperinsulinemic euglycemic clamp (Table 2). Changes in metabolic parameters pre to post intervention were estimated using a paired t-test. Pre and post intervention biopsies were used for gene expression analysis. Due to the problem that RNA isolation of all IMAT samples tissue material was not sufficient or RNA RIN values did not match quality requirements for gene expression analysis we were forced to remove all IMAT samples and used only the remaining muscle samples for gene expression analysis. Pre to post differential gene expression was estimated using one way ANOVA.

**Table 2.** Subject demographics: Longitudinal Intervention Study (N = 21)

|  | **<u>Pre-Intervention</u>** | **<u>Post-Intervention</u>** |
| --- | --- | --- |
| **<u>Age (yrs)</u>** | 46.5±2.2 |  |
| **<u>BMI (kg/m<sup>2</sup>)</u>** | 34.7±1.0 | 30.7±1.0 * |
| **<u>Body Weight (kg)</u>** | 96.9±2.7 | 85.9±2.6 * |
| **<u>RelFat (%)</u>** | 41.3±1.5 | 38.0±1.8 * |
| **<u>Fat free mass (kg)</u>** | 56.7±1.8 | 52.9±1.5 * |
| **<u>Fasting glucose (mg/dl)</u>** | 93.2±2.7 | 90.5±2.6 |
| **<u>VO<sub>2</sub>peak (L/min)</u>** | 2.2±0.1 | 2.5±0.1 * |
| **<u>Insulin Sensitivity (mg/kg/min)</u>** | 3.5±0.4 | 5.4±0.5 * |

Values are means±SEM. \* = significantly different from pre-intervention, p<0.05.

## Models and Statistics

To estimate the impact of gene expression on FG and GIR we utilized linear multivariate regression models. Thus we created a predictive model for each gene, simultaneously predicting clinical parameters based on gene expression in muscular and IMAT tissue. Models can be formalized in matrix notation as: **Y** = β**X**_**j**_ + ε, where **Y** is a matrix of the sampled response variables GIR (g) and FG (f) and **X** a matrix of the predictor values, the expression of gene j in muscle (m) and IMAT (i). β forms the 2×2 matrix of the four estimated regression coefficients β_mg_, β_mf_, β_ig_ and β_if_ describing the four relationships between tissue specific gene expression and response variables. The residues or errors are formed in ε. Our model can be interpreted as a mixture model that allows us to jointly estimate four coefficients to predict insulin sensitivity and glucose homeostasis from gene expression in muscle and IMAT. Genes that contributed the most to insulin sensitivity and glucose homeostasis were then scored based on log-likelihood and negative log-likelihood of the single regression models (S1). Analysis was done using Matlab R2020a.

Participant kNN-Networks were generated on cluster wise gene expression profiles with k = 3 nearest neighbors, using Euclidean distance metric. Predictive classification score was calculated as ratio of each participant direct neighbor’s clinical classification.

Further information, methods and protocols for genetic work and statistics are described in the Supplementary File S1.

## Results

### Multivariate regression unravels tissue specific gene expression patterns correlating with insulin resistance

We first compared demographic and metabolic parameter of participants. As expected, we found gender specific differences for RelFat (p<0.05), FFM (p < 0.001) and height (p<0.01) and for participants with obesity and T2D we found significant differences in GIR (p<0.05) and FG (p<0.001) (Table 1, Figure 1AB, S1A). Additionally, as expected, we found that low GIR values correlate with high FG levels (Figure S1B). As shown in Figure 1B, individuals classified as OB (BMI > 30 & FG < 125 md/dl) exhibited a wide range of insulin sensitivity (GIR 0.8 – 11.1 mg/kg/min). Some individuals with strong progressed insulin sensitivity (< 3 mg/kg/min) were still able to maintain FG levels below 125 mg/dl. In contrast, some diabetic individuals exhibited a better GIR with levels up to 4 mg/kg/min while being diagnosed as diabetic. To explore whether transcriptional changes in muscle and IMAT tissue at the transition from obesity with compensated insulin resistance (normoglycemia) to T2DM (hyperglycemia) may reflect the inconsistency between FG and GIR we performed a multivariate regression analysis to identify genes with a strong expressional link to insulin resistance and glucose homeostasis.

After multivariate regression analysis we selected the 59 top genes (Figure S1C) contributing to GIR and FG and used k-means to cluster them into three clusters of 14, 23 and 22 genes with distinct expression patterns in muscle and IMAT (Figure 1C, S1D, Table S1). We compared the gene cluster corresponding β values and observed three distinct patterns (Figure 1D, S1E): Cluster 1 (blue) combines genes whose expression is positively associated with GIR and negatively with FG in both tissues, muscle and IMAT, thus correlating with a healthy glucose metabolism. Cluster 2 contains genes that positively correlate with GIR and negatively with FG in muscle similarly to cluster 1, while no such effect was observed for these genes in IMAT. Cluster 3 to the contrary, contains genes with an opposing effect to GIR and FG exclusively in muscle while again no such impact was observed in IMAT.

Our results suggest that these largely different gene expression profiles of muscle and IMAT are associated with varying impact on glucose metabolism changes. In particular, expression of *PDK4*, which has been linked to diabetes and glucose metabolism previously [13], shows a high correlation with GIR and FG in muscle but almost none in IMAT (Figure 1D). We also identified genes with opposing effects on glucose metabolism in muscle compared to IMAT, such as *UBTD1* and *ST3GAL2*: Both genes show strong positive coefficients with FG and negative coefficients with GIR in muscle whereas in IMAT we observe negative β values with FG and positive with GIR (Figure 1D). This effect was mainly associated with cluster 3. In contrast, *NAPB* from cluster 2, shows exactly opposite associations. In a third observation we found genes, here represented by *SIN3A*, that seem to have a relatively high predictive value for FG but low or none for GIR in muscle but completely opposing values in IMAT (high on GIR low on FG).

Taken together, the here identified genes in three clusters show a striking association of muscle gene expression with GIR and FG, while only genes in cluster 1 also associate IMAT gene expression with glucose homeostasis and insulin sensitivity (Figure 1D, S1E). These results suggest that muscular gene expression profiles allow for a more specific and detailed characterization and classification of individual physiological states than serum based physiological parameters alone are able to.

### Gene expression based classification enables a refined view on the individual physiological state of obese patients

To test our hypothesis that gene expression patterns are superior in categorizing individual insulin resistance states compared to conventional clinical markers we performed a k-nearest-neighbor (kNN) classification for each tissue and gene cluster solely based on expression profiles. Thus, we generated six nearest neighbor networks (NNNs) representing expression based participant similarities for all 16 individuals (Figure 2A). Based on direct network neighbors we then calculated a predictive classification score for each individual (Figure 2A, S2). For muscle, we found for five of the 16 participants (two OB: Pb029, Pb043; three T2D: Pb034, PB053, Pb032) a non-unique classification over all three NNN. After averaging over the three clusters, one participant with obesity was classified as T2D and two participants with T2D were classified as OB.

**Figure 2:**
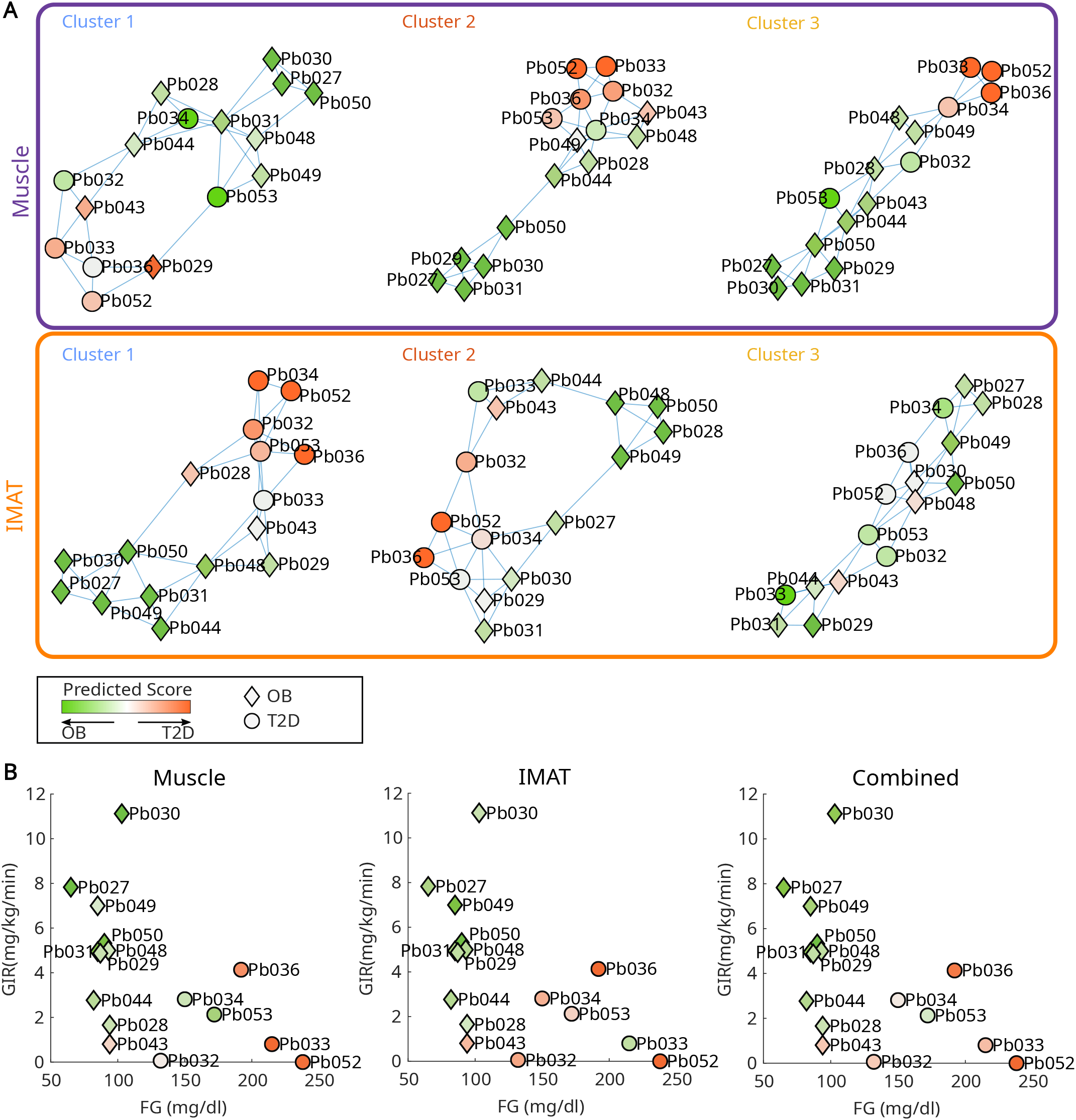
Gene expression based participant classification reveals a refined view on physiological states. **(A)** k-nearest-neighbor networks for the three cluster and two tissues respectively. Nodes refer to individual subjects. Node outer color refers to assigned clinical classification OB or T2D. Node inner color refers to estimated disease state based on connected individuals. (**B**) Scatter plot displays clinical parameter FG and GIR for all subjects. Node shape refers to assigned clinical classification OB (diamond) or T2D (dot). Node inner color refers to estimated disease state across all three gene cluster for muscle (left), IMAT (middle) and a combination of both tissues (right).

For the IMAT tissue derived NNNs we found for seven participants a predicted classification that differs from the clinical (three OB: Pb048, Pb028, Pb043; four T2D: Pb034, Pb053, Pb033, Pb032). IMAT gene expression averaged over the clusters classified two participants different form clinical classification: Pb043 (OB) and Pb033 (T2D). Averaging over both tissues, we identified two participants with divergent classification: Pb043 and Pb053. Over all, classification for OB classified participants was more consistent than for T2D with only two participants consistently classified as T2D (Pb036, Pb052).

When comparing NNN based classification to metabolic parameters, we found that the estimated probability of being predicted as T2D correlates with decreasing GIR rates (Figure 2B) for OB participants in both, muscle and IMAT tissue. In contrast, classification for hyperglycemic participants clinically classified as diabetic (FG > 125mg/dl), did not correlate with either GIR or FG in both tissues. These results suggest that beyond a binary clinical classification of T2D with FG levels > 125 mg/dl, there is a continuous development from insulin resistance to diabetes that follows individual traces with an individual diagnostic potential to predict a high or low risk for diabetes or the response interventions.

### ST3GAL2, SIN3A, ARF1 and AASS mRNA levels in muscle tissue predict intervention response

To evaluate whether gene expression profiles within muscle and/or IMAT define individual health states with predictive potential for disease progression or modulation of insulin sensitivity, we analyzed 17 human individuals with obesity, with and without prediabetes, undergoing a combined weight loss and exercise training intervention study (Table 2). Clinical parameters such as GIR, FG, BW, RelFat, FFM and BMI were measured pre and post intervention. Almost all individuals showed an increase of GIR (p = 2.2*10^−5^) after the intervention and a decrease of BMI (p = 2.3*10^−8^), BW (p = 6.1*10^−8^), RelFat (p = 2.7*10^−6^) and FFM (p = 1.4*10^−6^). A general change of FG levels upon intervention could not be observed (p = 0.12) (Figure 3A). However, when correlating the relative pre/post change (Δ%) between all clinical parameters we found, a change of GIR did only significantly correlate with a change in BW (Figure S3). A decrease of FG in turn, was significantly correlated with a relative decrease in BMI, FFM and BW.

**Figure 3:**
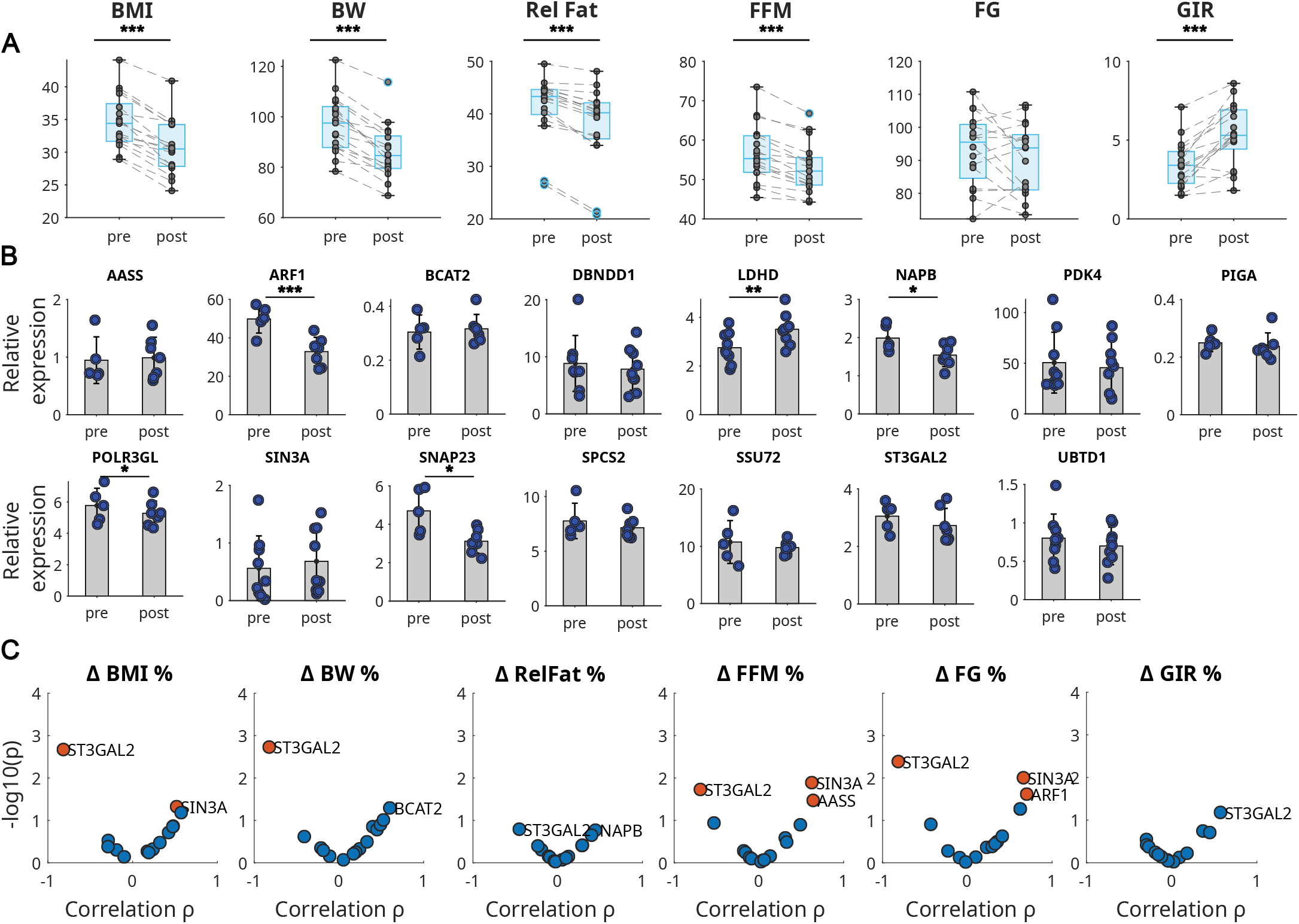
ST3GAl2, SIN3A, ARF1 and AASS mRNA expression in muscle predict intervention response. **(A)** Clinical parameters pre and post intervention. Significance levels refer to paired t test. **(B)** mRNA levels of selected genes pre and post intervention. Significant differences in pre/post expression was estimated using 1 way ANOVA. **(C)** Correlation volcano plots for pre-intervention gene expression with relative change of clinical parameters between pre and post intervention. Significantly correlated mRNAs are colored orange.

Subsequently, to test if these physiological changes are linked to individual gene expression in muscle, biopsies taken before and after intervention were used for RNA expression analysis. Six pre and eight post intervention muscle samples did not match quality requirements and were removed from subsequent analyses. Out of the previously selected 59 top genes initially identified in our first patient cohort, we combined various criteria to select 15 candidate genes from all clusters for validation. Gene expression in muscle tissue was measured with quantitative RT-PCR (Table S1) within this second independent intervention trial. Among those *SIN3A, UBTD1, ST3GAL2* and *NAPB* showed notable β value profiles (Figure 1D). *AASS, DBNDD1, PDK4, PIGA, POLR3GL, SNAP23, SPCS2, SSU72* and *UBTD1* could be linked to diabetes associated SNPs identified with the TD2M portal [14] and *ARF1, BCAT2* and *LDHD* could be linked to skeletal muscle lipid metabolism and insulin resistance [15-17]. *PDK4* was included as a well described marker for muscle insulin resistance and as a potential therapeutic target [18]. From these 15 genes, five (*LDHD, ARF1 NAPB, POLR3GL* and *SNAP23*) showed a significant change in expression between pre and post intervention (Figure 3B). Since we hypothesized that distinct gene expression states may refer to individual disease states, we tested selected genes on their predictive potential for individual intervention response. To this end, we correlated individual pre-intervention gene expression with the relative change (pre to post) of the clinical parameters BMI, BW, RelFat, FFM, FG and GIR (Figure 3C). ΔGIR and ΔRelFat mass could not be significantly with any of the genes tested. A change of the remaining parameters could be significantly predicted by the genes *ST3GAL2* (FG, BW, FFM and BMI), *SIN3A* (FG, FFM and BMI), *ARF1* (FG) and *AASS* (FFM) (Figure 1C, S4, Table S2). In contrast to the five genes that change expression after intervention (*LDHD, ARF1, NAPB, POLL3GL, SNAP23*), none of the four genes identified with predictive character appeared to be differentially expressed between pre- and post-intervention (Figure 3B). Together, these findings indicate that individual susceptibility to exercise intervention for the improvement of glucose homeostasis is independent of the individual clinical parameters, but correlates with individual gene expression profiles prior to intervention. We next compared these four identified genes with *PDK4*, a well-described muscle marker for insulin resistance. To our surprise, *PDK4* was not significant associated with any intervention-induced change of metabolic parameters (Figure S4).

Next, we found that low expression levels of three out of the four genes (*AASS, ARF1* and *SIN3A*) were associated with a good health prognosis. In particular, *ARF1* showed a significant decrease of expression upon exercise intervention. In turn, *ST3GAL2* was the only gene for which increased expression levels in muscle tissue increases likeliness of an effective intervention. In summary, within this independent intervention trial we were able to validate our proposed predictive potential of muscle gene expression profiles for individual insulin resistance states.

## Discussion

In this work, we showed that human transcriptional profiles of muscle and IMAT tissue from obese and diabetic individuals are differentially coupled to insulin resistance and glucose homeostasis. We identified predictive gene clusters that mirror genetic states reflecting a continuous progression from early insulin resistance to T2D following individual traits. From a subset, we identified the genes *AASS, ARF1, SIN3A* and *ST3GAL2* to predict individual exercise intervention response to improve impaired glucose metabolism.

We started our analysis with the observation that there is a wide range of GIR measurements that overlap between OB and T2D. We hypothesized the binary clinical classification of T2D does not reflect individual underlying genetics and that specific gene expression patterns of skeletal muscle and/or IMAT tissue might have the potential to identify and predict individuals with a high or low risk to develop diabetes or to predict individual susceptibility to interventions.

Our multivariate regression analysis, revealed a strong association of muscle to GIR and FG for all three gene clusters, while IMAT was associated with glucose homeostasis and insulin resistance only with the genes in cluster 1. Together with the observation that estimates for β coefficients of cluster 2 were contrary in muscle and IMAT we concluded that IMAT and muscle contribute differentially to glucose metabolism. However, we see higher variance in IMAT gene expression [12] which might also weaken a detectable correlation. Reason’s for IMAT’s increased variability may arise from technical difficulties to dissect IMAT from muscle resulting in less material for RNA extraction or a higher heterogeneity in IMAT tissue itself being composed of multiple cell types such as pre-adipocytes, adipocytes, adipocyte-like cells, myoblasts, stromal and vascular cells. Participant classification based on kNN-networks revealed that insulin sensitivity could be accurately predicted for non-diabetic subjects from gene expression patterns, whereas hyperglycemic subjects were scored differently from clinical classification in several cases. These observations are consistent with hyperglycemia usually being a consequence of pancreatic beta-cell failure, and insulin sensitivity is associated with multiple organ malfunctions and, in particular, the skeletal muscle as the primary organ for glucose uptake [19]. Although GIR measured with hyperinsulinemic-euglycemic clamp, is still the gold standard to directly measure insulin resistance it is highly invasive and time consuming and with very limited predictive potential. FG levels by themselves are unlikely to identify pre-diabetic or obese individuals with impaired insulin sensitivity, but rather individuals at late stages with an increased risk for irreversible damages of tissues and organs [20]. We thus conclude, that both parameters are not suitable for reliable for early diagnosis and disease progression prognosis. In contrast, with the here identified gene expression profiles that only represent a muscle specific state of individual insulin resistance, we could identify their predictive potential for the characterization of the individual insulin sensitivity.

This predictive potential was finally tested on muscle tissue from an additional independent cohort of 18 individuals with impaired glucose metabolism undergoing 12 weeks of combined weight loss and exercise training. By correlating the pre-intervention expression levels of our candidate genes with the relative change of the clinical parameters upon intervention we identified 4 genes with significant predictive value: *AASS, ARF1, SIN3A* and *ST3GAL2. AASS, ARF1* and *SIN3A* indicated a positive prognosis with a lower level of expression. *AASS* encodes for the enzyme *Aminoadipate-Semialdehyde Synthase* that is involved in mammalian lysine degradation and in hyperlysinemia [21], but was not described in the context of impaired glucose metabolism, insulin resistance or diabetes yet. Beside its predictive potential we also found that ADP Ribosylation Factor 1 (*ARF1*) expression was significantly reduced after intervention. The Ink4/Arf locus was recently linked to insulin resistance in mice [22] and ADP-ribosylation factor was linked to insulin signaling in *Drosophila* [23]. The transcription factor SIN3 Transcription Regulator Family Member A (*SIN3A)* was recently linked to glucose metabolism in murine β-cells [24]. It could be further shown that *Sin3a* is an insulin sensitive FOXO1 corepressor of glucokinase in murine livers [25]. Very recently, *SIN3A* was shown to negatively regulate insulin receptor *(Insr)* mRNA in mice and human muscle [26]. Finally, ST3 Beta-Galactoside Alpha-2,3-Sialyltransferase 2 (*ST3GAL2*) was the only gene that was identified to positively predict exercise response with a high expression. Mice lacking the ST3Gal-II protein, have been shown to develop obesity and insulin resistance after 7-9 month of age [27]. In summary, 3 of the 4 predictive genes identified by us have already been linked to insulin resistance and diabetes but their predictive potential has not been explored yet.

We herein identified novel markers to predict impaired insulin sensitivity in the human muscle and found four markers that predict individual exercise intervention response in diabetic patients. These findings may help to classify and characterize obese, pre-diabetic or diabetic individuals more precisely than using FG alone. Clamp GIR measurements are highly invasive and may be partly replaced by use of our marker genes. Additionally we anticipate these findings may also help to develop precise and individualized interventions strategies for diabetic and obese risk patients.

## Data Availability

All data produced in the present study are available upon reasonable request to the authors

## Contributors

DL, BCB and SH conceived the research question and designed and planned the study. DL, SS, MW, AK, LP, DEK, AW and MK prepared the data. DL analyzed the data. DL, BCB and SH wrote the manuscript. All authors critically revised the manuscript. DL had full access to all the data in the study and had final responsibility to submit for publication. BCB and SH had access to and verified the study data.

## Data sharing

Gene expression profiles and clinical data can be shared upon request to BCB. Statistical code as not provided in the manuscript or supplementary information is available upon request to DL

## Acknowledgements

We gratefully thank Ricardo Berutti for bioinformatics support provided at the Institute of Human Genetics at the Helmholtz Zentrum München and Shohreh Esmaeili for technical assistance.

