## Supplementary Information for "Skeletal muscle and intermuscular adipose tissue gene expression profiling identifies new biomarkers with prognostic significance for insulin resistance progression and intervention response"

**Figure S1**

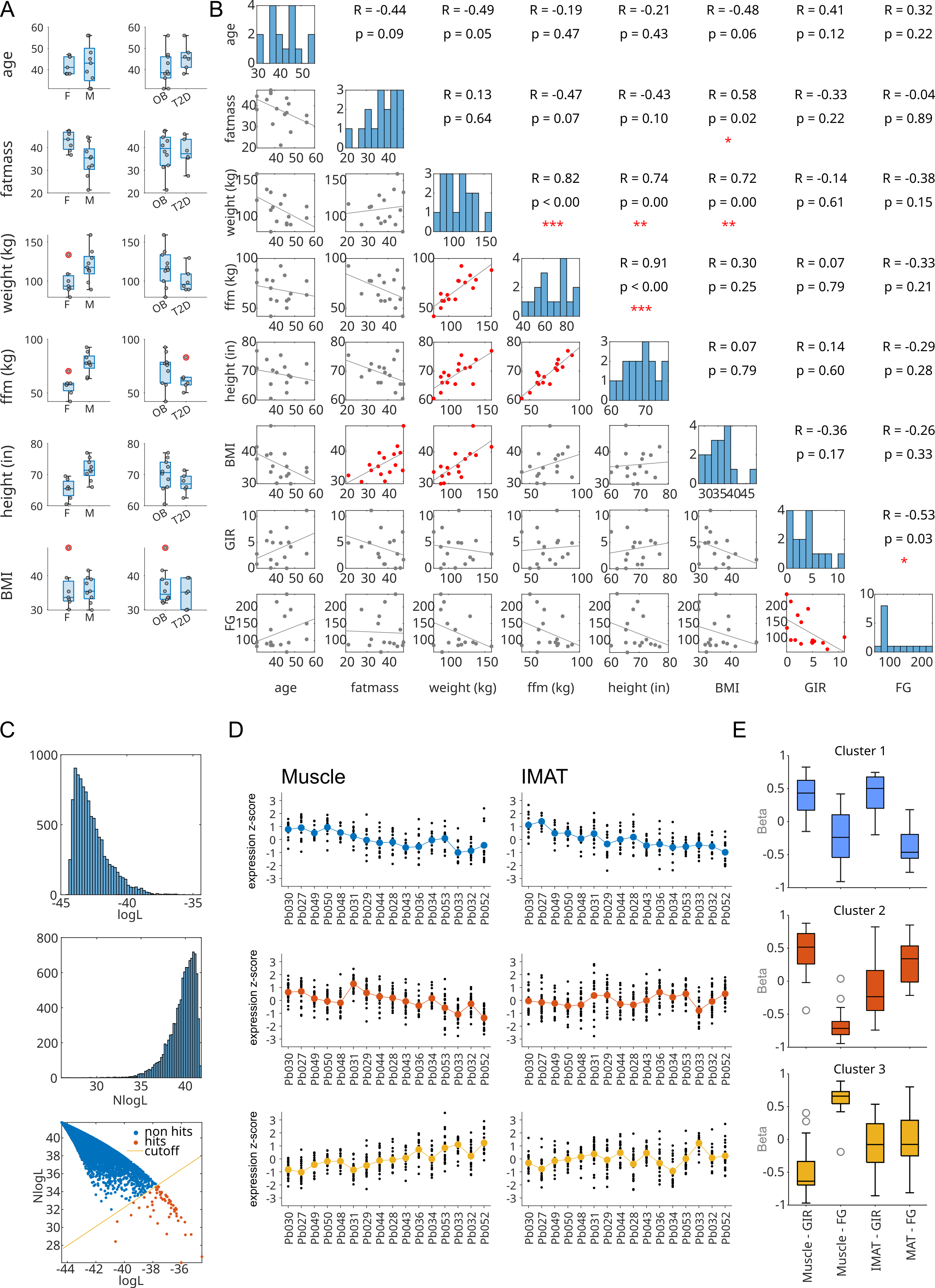

**Figure S1**

**A)** Boxplot comparing clinical parameter distributions between subjects of different classifications and gender. OB: obese, T2D: type 2 diabetic F: female, M: male. Red marked dots indicate outliers.

**B)** Correlation plot for pairwise comparisons between measured clinical parameter including histograms showing parameter distribution. Red dots indicate significant correlation. Upper right triangle lists correlation coefficients and p-values.

**C)** Distribution of mRNA wise estimated regression model log-Likelihood (top) and Negative log-likelihood (middle) and scattered versus each other (bottom). Yellow cutoff line indicates selection function, red dots refer to mRNAs selected for further investigation.

**D)** Box plots show distribution of the four regression coefficients for each cluster. Horizontal lines refer to median, boxes to upper and lower quartile and whiskers to maxima and minima of the distributions. Dots denote outliers.

**E)** Subject wise normalized mRNA expression for the three gene clusters for Muscle (left) and IMAT (tissue).

**Figure S2**

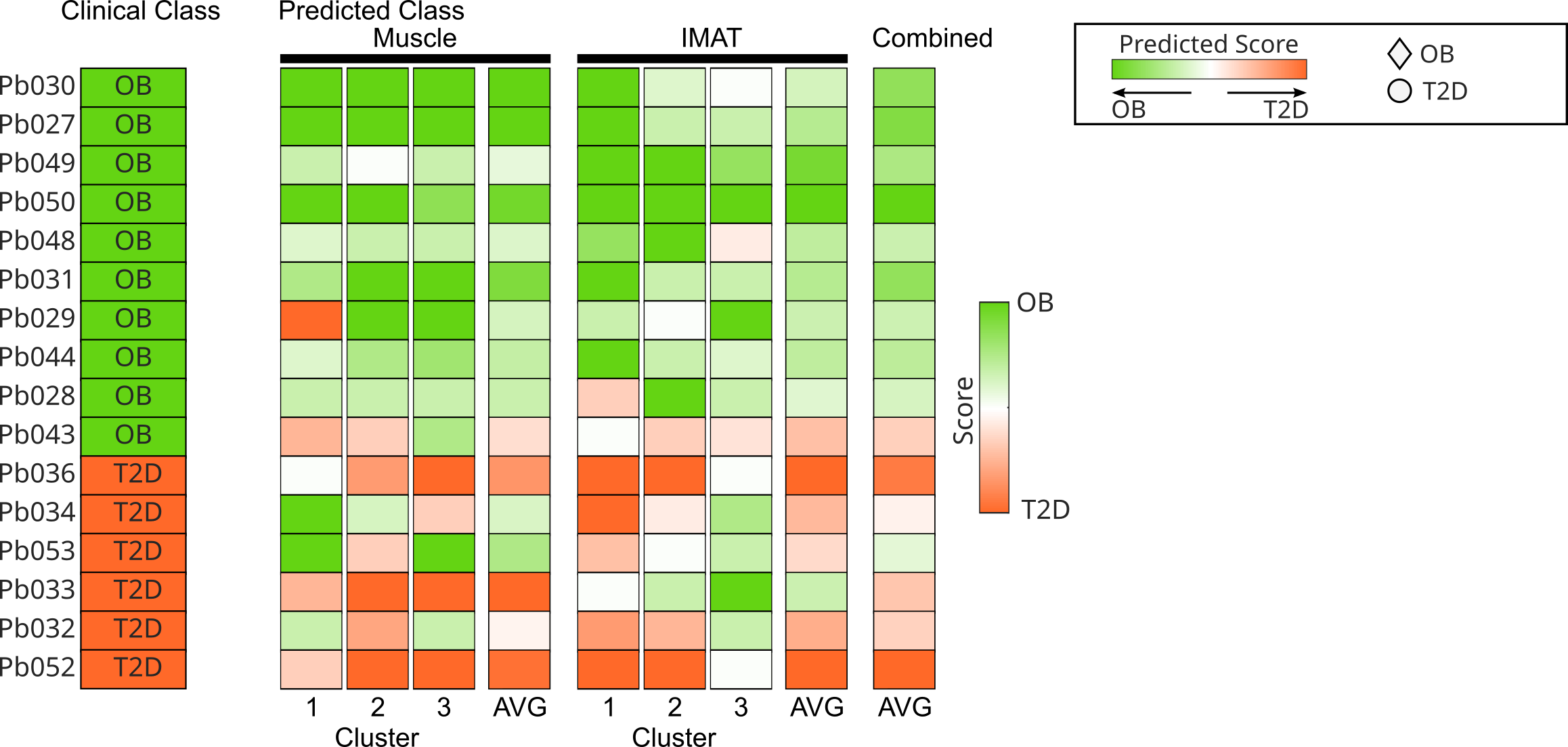

**Figure S2**

Estimated classification scores by k-nearest neighbor classification for each cluster and tissue including average scores for each tissue and for both tissues combined.

Figure S3

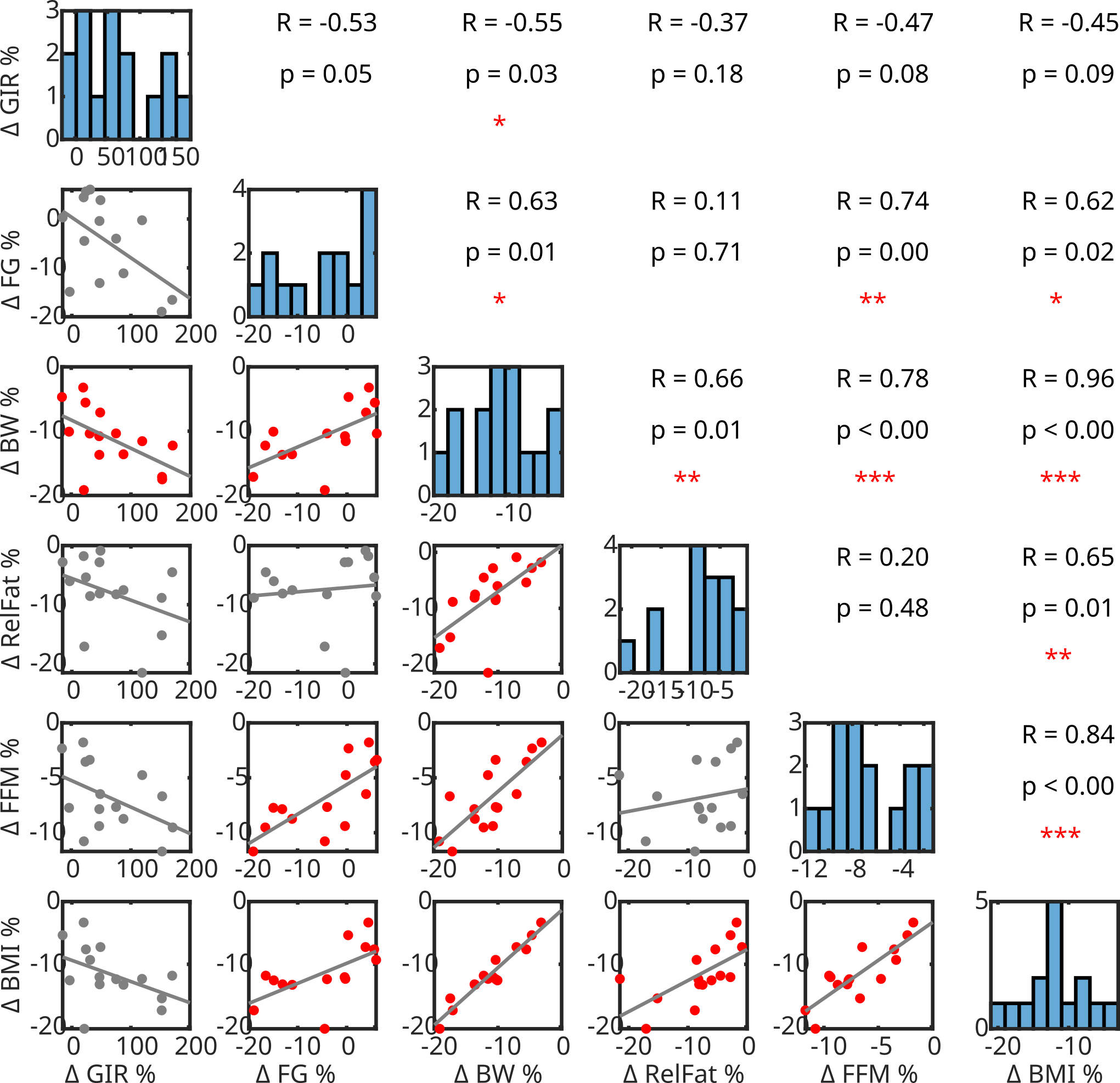

**Figure S3**

Correlation plot for pairwise comparisons between with relative change (pre to post) of clinical parameter including histograms showing parameter distribution. Red dots indicate significant correlation. Upper right triangle lists correlation coefficients and p-values.

**Figure S4**

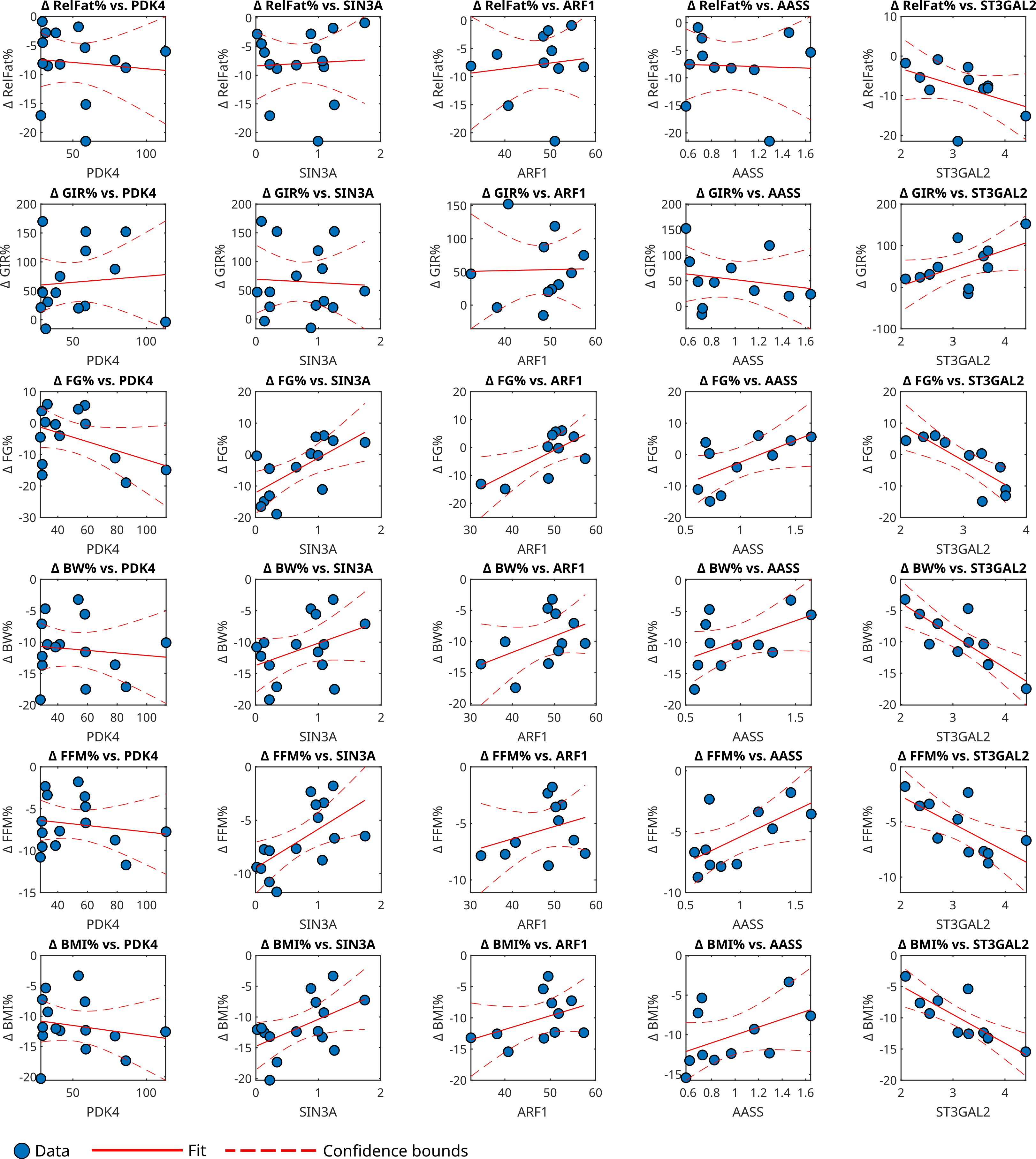

**Figure S4**

Correlation plots for five selected mRNAs with clinical parameters. Red lines refer to estimated correlation, dashed lines to confidence intervals.

**Table S1**

| Symbol | logL | NlogL | β muscle - GIR | β muscle - FG | β IMAT - GIR | β IMAT - FG | Cluster |
| --- | --- | --- | --- | --- | --- | --- | --- |
| AASS | -35.75 | 30.59 | 0.77 | -0.88 | 0.01 | 0.35 | 2 |
| ADAR | -38.20 | 33.88 | 0.71 | -0.45 | 0.23 | -0.02 | 2 |
| ALDH6A1 | -37.65 | 34.59 | 0.21 | -0.23 | 0.52 | -0.46 | 1 |
| AMFR | -38.38 | 32.30 | -0.33 | 0.82 | -0.12 | -0.09 | 3 |
| ARF1 | -36.85 | 33.08 | -0.69 | 0.76 | 0.46 | -0.48 | 3 |
| BCAT2 | -36.57 | 32.46 | 0.63 | -0.44 | 0.20 | -0.47 | 1 |
| BDH1 | -34.56 | 26.73 | 0.68 | -0.30 | 0.22 | -0.56 | 1 |
| C7orf25 | -37.59 | 34.51 | -0.66 | 0.64 | 0.24 | -0.22 | 3 |
| CCDC176 | -37.31 | 33.13 | 0.38 | -0.77 | 0.41 | 0.18 | 1 |
| CDK7 | -38.96 | 32.19 | 0.28 | -0.77 | -0.30 | -0.02 | 2 |
| CLEC3B | -36.91 | 32.72 | 0.61 | -0.80 | 0.21 | -0.21 | 2 |
| DBNDD1 | -36.50 | 31.99 | -0.68 | 0.89 | 0.05 | -0.26 | 3 |
| DDR1 | -37.72 | 33.85 | 0.83 | -0.02 | -0.20 | -0.65 | 1 |
| DHCR24 | -39.53 | 31.36 | -0.01 | 0.34 | 0.73 | -0.17 | 1 |
| DHTKD1 | -37.79 | 34.50 | 0.61 | -0.59 | -0.03 | -0.19 | 1 |
| ENPP1 | -37.74 | 34.19 | 0.73 | -0.54 | -0.13 | 0.09 | 1 |
| FANCE | -37.02 | 29.26 | 0.55 | 0.10 | 0.54 | -0.54 | 1 |
| FRZB | -40.50 | 30.65 | 0.10 | 0.52 | -0.08 | 0.38 | 3 |
| GAB3 | -36.33 | 31.26 | 0.63 | -0.81 | -0.04 | -0.14 | 2 |
| GNG5 | -37.68 | 34.24 | -0.41 | 0.62 | -0.49 | 0.16 | 3 |
| GPT | -35.31 | 29.21 | 0.17 | 0.00 | 0.68 | -0.77 | 1 |
| HERC6 | -36.02 | 31.10 | 0.70 | -0.72 | 0.39 | -0.01 | 2 |
| HFE2 | -36.76 | 32.91 | -0.70 | 0.73 | 0.03 | 0.02 | 3 |
| HNRNPU-AS1 | -37.95 | 31.90 | 0.41 | -0.91 | -0.36 | 0.39 | 2 |
| HSPA2 | -36.40 | 32.15 | -0.73 | 0.76 | 0.05 | -0.06 | 3 |
| LDHD | -36.46 | 31.89 | 0.49 | -0.24 | 0.49 | -0.53 | 1 |
| MSL3 | -36.75 | 31.50 | 0.28 | 0.42 | -0.79 | 0.43 | 3 |
| NAPB | -36.35 | 31.55 | 0.53 | -0.55 | -0.71 | 0.46 | 2 |
| NEGR1 | -37.38 | 33.94 | 0.73 | -0.80 | -0.46 | 0.68 | 2 |
| PDK4 | -37.19 | 33.15 | -0.97 | 0.60 | 0.27 | -0.01 | 3 |
| PIGA | -35.72 | 29.17 | 0.87 | -0.60 | -0.14 | 0.55 | 2 |
| PITHD1 | -37.07 | 33.50 | -0.39 | 0.55 | -0.40 | 0.29 | 3 |
| POLR1E | -40.62 | 28.42 | -0.44 | 0.04 | 0.82 | 0.14 | 2 |
| POLR3GL | -36.49 | 31.79 | -0.61 | 0.80 | -0.16 | -0.25 | 3 |
| PPM1K | -36.34 | 32.01 | 0.85 | -0.83 | -0.23 | 0.23 | 2 |
| QSOX2 | -37.13 | 32.97 | 0.88 | -0.71 | -0.25 | 0.49 | 2 |
| RCAN1 | -37.06 | 33.50 | 0.40 | -0.19 | -0.86 | 0.80 | 3 |
| SAYSD1 | -37.42 | 33.32 | -0.32 | 0.46 | -0.28 | 0.40 | 3 |
| SF3A1 | -37.51 | 33.96 | -0.15 | 0.35 | 0.75 | -0.56 | 1 |
| SIN3A | -37.67 | 30.44 | -0.02 | -0.64 | 0.70 | -0.01 | 2 |
| SLFNL1-AS1 | -38.03 | 32.51 | 0.06 | 0.42 | 0.73 | -0.45 | 1 |
| SNAP23 | -36.99 | 33.42 | 0.76 | -0.75 | -0.37 | 0.34 | 2 |
| SPCS2 | -37.10 | 33.63 | -0.69 | 0.68 | -0.08 | 0.09 | 3 |
| SSBP2 | -37.75 | 34.21 | -0.37 | 0.42 | -0.39 | 0.59 | 3 |
| SSU72 | -36.71 | 32.58 | -0.69 | 0.70 | -0.13 | -0.25 | 3 |
| ST3GAL2 | -36.22 | 30.74 | -0.91 | 0.67 | 0.30 | -0.43 | 3 |
| SYAP1 | -37.40 | 33.83 | -0.86 | 0.65 | 0.54 | -0.62 | 3 |
| TAF1 | -37.81 | 33.30 | 0.25 | -0.71 | 0.47 | -0.05 | 2 |
| TAF11 | -38.05 | 32.86 | 0.33 | -0.94 | -0.59 | 0.85 | 2 |
| TIGD7 | -37.75 | 34.19 | 0.03 | -0.27 | -0.74 | 0.54 | 2 |
| TRA2A | -38.17 | 31.20 | 0.13 | -0.70 | -0.38 | 0.53 | 2 |
| TSPAN17 | -39.36 | 32.76 | -0.21 | 0.70 | -0.35 | -0.15 | 3 |
| TTC7B | -37.48 | 33.64 | -0.59 | 0.68 | -0.05 | -0.24 | 3 |
| UBTD1 | -36.72 | 32.18 | -0.67 | 0.65 | 0.47 | -0.81 | 3 |
| ZNF10 | -38.60 | 33.36 | 0.38 | -0.63 | -0.12 | 0.54 | 2 |
| ZNF561 | -36.86 | 31.15 | 0.52 | -0.85 | -0.23 | 0.23 | 2 |
| ZNF692 | -37.66 | 34.56 | 0.23 | -0.40 | -0.72 | 0.77 | 2 |
| ZNF75A | -36.84 | 32.83 | 0.36 | -0.66 | -0.47 | 0.21 | 2 |
| ZRANB2-AS1 | -36.01 | 26.06 | 0.33 | -0.91 | 0.59 | -0.32 | 1 |

**Table S1**

Top 59 Genes with high associations to GIR and FG. Log-likelihood, neg-log-Likelihood, β values and Clustering coefficients are given. Genes selected for qPCR validation with intervention cohort are color shaded.

| _Gene_ | _p RelFat%_ | _padj RelFat%_ | _coef RelFat%_ | _R2 RelFat%_ | _p GIR%_ | _padj GIR%_ | _coef GIR%_ | _R2 GIR%_ | _p FG%_ | _padj FG%_ | _coef FG%_ | _R2 FG%_ | _p BW%_ | _padj BW%_ | _coef BW%_ | _R2 BW%_ | _p FFM%_ | _padj FFM%_ | _coef FFM%_ | _R2 FFM%_ | _p BMI%_ | _padj BMI%_ | _coef BMI%_ | _R2 BMI%_ |
| --- | --- | --- | --- | --- | --- | --- | --- | --- | --- | --- | --- | --- | --- | --- | --- | --- | --- | --- | --- | --- | --- | --- | --- | --- |
| _LDHD_ | _0.495_ | _0.928_ | _-2.23_ | _0.037_ | _0.284_ | _0.949_ | _-34.66_ | _0.088_ | _0.735_ | _0.809_ | _1.67_ | _0.010_ | _0.447_ | _0.696_ | _-1.98_ | _0.045_ | _0.523_ | _0.944_ | _-1.09_ | _0.032_ | _0.298_ | _0.615_ | _-2.54_ | _0.083_ |
| _UBTD1_ | _0.701_ | _0.944_ | _-2.11_ | _0.012_ | _0.866_ | _0.949_ | _-9.34_ | _0.002_ | _0.433_ | _0.590_ | _6.99_ | _0.052_ | _0.851_ | _0.851_ | _0.82_ | _0.003_ | _0.259_ | _0.647_ | _3.17_ | _0.097_ | _0.536_ | _0.615_ | _2.56_ | _0.030_ |
| _PDK4_ | _0.741_ | _0.944_ | _-0.02_ | _0.009_ | _0.748_ | _0.949_ | _0.21_ | _0.008_ | _0.124_ | _0.310_ | _-0.15_ | _0.186_ | _0.699_ | _0.749_ | _-0.02_ | _0.012_ | _0.573_ | _0.944_ | _-0.02_ | _0.025_ | _0.498_ | _0.615_ | _-0.03_ | _0.036_ |
| _DBNDD1_ | _0.400_ | _0.858_ | _-0.28_ | _0.055_ | _0.180_ | _0.949_ | _4.38_ | _0.134_ | _0.755_ | _0.809_ | _-0.21_ | _0.008_ | _0.510_ | _0.696_ | _-0.18_ | _0.034_ | _0.885_ | _0.944_ | _0.03_ | _0.002_ | _0.723_ | _0.723_ | _-0.09_ | _0.010_ |
| _SIN3A_ | _0.851_ | _0.944_ | _0.59_ | _0.003_ | _0.853_ | _0.949_ | _-5.87_ | _0.003_ | **_0.010_** | _0.074_ | _11.05_ | _0.439_ | _0.141_ | _0.414_ | _3.55_ | _0.159_ | **_0.013_** | _0.140_ | _3.64_ | _0.390_ | **_0.047_** | _0.329_ | _4.38_ | _0.270_ |
| _SPCS2_ | _0.385_ | _0.858_ | _1.01_ | _0.085_ | _0.380_ | _0.949_ | _-8.70_ | _0.087_ | _0.958_ | _0.958_ | _-0.12_ | _0.000_ | _0.470_ | _0.696_ | _0.61_ | _0.060_ | _0.748_ | _0.944_ | _-0.16_ | _0.012_ | _0.487_ | _0.615_ | _0.52_ | _0.055_ |
| _ARF1_ | _0.711_ | _0.944_ | _0.10_ | _0.016_ | _0.949_ | _0.949_ | _0.15_ | _0.000_ | **_0.024_** | _0.120_ | _0.76_ | _0.491_ | _0.166_ | _0.414_ | _0.26_ | _0.202_ | _0.326_ | _0.698_ | _0.11_ | _0.107_ | _0.194_ | _0.485_ | _0.22_ | _0.180_ |
| _BCAT2_ | _0.393_ | _0.858_ | _25.17_ | _0.082_ | _0.423_ | _0.949_ | _-202.04_ | _0.073_ | _0.054_ | _0.162_ | _75.96_ | _0.389_ | _0.051_ | _0.382_ | _37.97_ | _0.360_ | _0.127_ | _0.381_ | _17.64_ | _0.239_ | _0.066_ | _0.329_ | _32.11_ | _0.328_ |
| _NAPB_ | _0.170_ | _0.858_ | _8.98_ | _0.198_ | _0.902_ | _0.949_ | _-7.29_ | _0.002_ | _0.413_ | _0.590_ | _7.56_ | _0.085_ | _0.560_ | _0.700_ | _2.87_ | _0.039_ | _0.800_ | _0.944_ | _-0.72_ | _0.007_ | _0.473_ | _0.615_ | _3.12_ | _0.059_ |
| _PIGA_ | _0.224_ | _0.858_ | _86.00_ | _0.160_ | _0.724_ | _0.949_ | _-221.11_ | _0.014_ | _0.317_ | _0.590_ | _100.37_ | _0.125_ | _0.126_ | _0.414_ | _75.90_ | _0.240_ | _0.695_ | _0.944_ | _11.82_ | _0.018_ | _0.137_ | _0.421_ | _65.73_ | _0.229_ |
| _AASS_ | _0.908_ | _0.944_ | _-0.65_ | _0.002_ | _0.570_ | _0.949_ | _-27.05_ | _0.037_ | _0.054_ | _0.162_ | _13.91_ | _0.390_ | _0.097_ | _0.414_ | _6.21_ | _0.275_ | **_0.034_** | _0.169_ | _4.32_ | _0.410_ | _0.140_ | _0.421_ | _4.98_ | _0.225_ |
| _ST3GAL2_ | _0.161_ | _0.858_ | _-3.99_ | _0.206_ | _0.066_ | _0.949_ | _43.03_ | _0.327_ | **_0.004_** | _0.061_ | _-11.45_ | _0.664_ | **_0.002_** | **_0.028_** | _-5.22_ | _0.677_ | **_0.019_** | _0.140_ | _-2.50_ | _0.477_ | **_0.002_** | **_0.032_** | _-4.60_ | _0.668_ |
| _POLR3GL_ | _0.944_ | _0.944_ | _-0.19_ | _0.001_ | _0.196_ | _0.949_ | _23.40_ | _0.200_ | _0.526_ | _0.657_ | _-2.32_ | _0.052_ | _0.241_ | _0.516_ | _-1.90_ | _0.167_ | _0.115_ | _0.381_ | _-1.72_ | _0.281_ | _0.416_ | _0.615_ | _-1.32_ | _0.084_ |
| _SSU72_ | _0.774_ | _0.944_ | _0.22_ | _0.010_ | _0.597_ | _0.949_ | _3.38_ | _0.032_ | _0.355_ | _0.590_ | _0.97_ | _0.107_ | _0.615_ | _0.709_ | _0.27_ | _0.029_ | _0.944_ | _0.944_ | _0.02_ | _0.001_ | _0.574_ | _0.615_ | _0.27_ | _0.036_ |
| _SNAP23_ | _0.389_ | _0.858_ | _1.46_ | _0.084_ | _0.641_ | _0.949_ | _-6.81_ | _0.025_ | _0.236_ | _0.506_ | _3.45_ | _0.170_ | _0.320_ | _0.601_ | _1.20_ | _0.109_ | _0.855_ | _0.944_ | _0.13_ | _0.004_ | _0.334_ | _0.615_ | _1.04_ | _0.104_ |

**Table S2**

Statistics of regression models predicting relative change of clinical parameters from pre intervention gene expression. P-values, adjusted p-values, coefficients and R^2^ are summarized for each gene and model.

**Table S3**

**Primers used for qPCR:**

AASS forward	5’-GAATCGGCGGGCCATTCAT-3‘,

AASS reverse	5’-CTGAGCTTTTATTGTGTGGGAGA-3’;

ARF1 forward	5’-ATGGGGAACATCTTCGCCAAC-3‘,

ARF1 reverse	5’-GTGGTCACGATCTCACCCAG-3’;

BCAT2 forward	5’-GCTCAACATGGACCGGATG-3‘,

BCAT2 reverse	5’-CCGCACATAGAGGCTGGTG-3’;

DBNDD1 forward	5’-TCTTTGCTGACTCGGACGAC-3‘,

DBNDD1 reverse	5’-CCACAGTGAGAAACGTGTCCA-3’;

LDHD forward	5’-CCGTAGCCCGCATTGAGTT-3‘,

LDHD reverse	5’-CTGCTGGACTATCTCCTCTGT-3’;

NAPB forward	5’-CAGGAAACGCATTTTGTCAGG-3‘,

NAPB reverse	5’-GGGATCTGCCTTTTTGTAAGCAT-3’;

PDK4 forward	5’-GGAAGCATTGATCCTAACTGTGA-3‘,

PDK4 reverse	5’-GGTGAGAAGGAACATACACGATG-3’;

PIGA forward	5’-GTTGGCAGTTTTCAACTTCCTCT-3‘,

PIGA reverse	5’-TGGCCCAGTGGCATCTATTG-3’;

POLR3GL forward	5’-ATCAGATGTCAGGTCCGATTGA-3‘,

POLR3GL reverse	5’-AGCAGAATTGTAATCCGTTCCTT-3’;

SIN3A forward	5’-ATGAGTCTCTGGAAAGTACGAGG-3‘,

SIN3A reverse	5’-GCATCTGGTAGGAATTGTCCAAA-3’;

SNAP23 forward	5’-CCTGTGGAGTTTAATCATGCCA-3‘,

SNAP23 reverse	5’- CCACAGCATTTGTTGAGTTCTG -3’;

SPCS2 forward	5’-CGTAGCGGCTTGTTGGATAAG-3‘,

SPCS2 reverse	5’-GTGAGGCGACCATCAATTAGAC-3’;

SSU72 forward	5’-TCCCGACAAGCCCAATGTTTA-3‘,

SSU72 reverse	5’-CTCTTCGCAAGTGAGGATCAG-3’;

ST3GAL2 forward	5’-CGTCTGGACCCGAGAGAAC-3‘,

ST3GAL2 reverse	5’-GCCAGGCACTATCTGGAACA-3’;

TBP forward	5’-AACAACAGCCTGCCACCTTA-3‘,

TBP reverse	5’-GCCATAAGGCATCATTGGAC-3’;

UBTD1 forward	5’-CGGAGCAAACGGGATGAGTT-3‘,

UBTD1 reverse	5’-GCGGTGACAGGCAGTAGAT-3’.

**Supplementary Information and Methods**

**Cohort Ethnicity**

The human transcriptional profiling dataset included 13 participants of Caucasian and three participants of Hispanic ethnicity. The longitudinal intervention study included 12 individuals of Caucasian ethnicity, three Hispanic, one East Indian, and one African American. Ethnicity was not taken into account for statistical analyses.

**Longitudinal Intervention Study**

**Metabolic study:** Energy requirement was estimated based on DEXA determined fat free mass with the macronutrient intake at 55% carbohydrate, 15% protein, and 30% fat. Volunteers spent the night on the CTRC to ensure compliance with the overnight fast. The metabolic study consisted of a basal muscle biopsy followed by a 3-hour hyperinsulinemic euglycemic clamp at 40 mU/m^2^/min for 3 hours. A variable infusion of 20% dextrose was infused to maintain blood glucose ~90 mg/dl.

**12 week intervention program:** Everyone received a low-calorie diet consisting of a meal replacement product that can be consumed as a liquid or made into a variety of food forms (Health Nutrition Technology Inc., Carmel California). Subjects were provided powdered HealthOne formula and instructed to consume 5 portions per day providing 890 kcal/d, 75 g or protein, 15 g fat and 110 g of carbohydrate and 100% of the DRI of all vitamins, minerals and micronutrients. Subjects were allowed to consume non-caloric beverages but no other food intake was allowed. The delivery of this weight loss program has been supervised by the Clinical Core of the NORC (P30 DK048520) at the Anschutz Health and Wellness Center on the campus of the Anschutz Medical Center. Subjects were seen weekly in one-on-one sessions with a registered dietician for help with the diet and to receive nutritional counseling.

Participants also underwent supervised endurance exercise training using well-described procedures used by the NORC energy balance core. Volunteers were asked to attend 4 sessions per week. Each session lasted 60 min and included a short warm-up of stretching exercises and walking, 40 to 50 min of endurance exercise, and a cool-down period. The exercise program consisted primarily of brisk walking or jogging, and was supplemented with rowing, stepping, or elliptical exercise to provide variety and relieve joint discomfort when necessary.

**2 week weight maintenance diet:** Three servings of meal replacement per day along with one meal of typical foods for one additional month to stabilize weight. This period of the dietary intervention was supervised by research dieticians who have extensive experience helping subjects maintain a reduced state for metabolic studies at the University of Colorado CTRC.

**Gene expression analysis.**

Total RNA was isolated using RNeasy Kit (Qiagen, Hilden, Germany) according to manufacturer’s instructions. cDNA synthesis was performed with QuantiTect Reverse Transcription Kit (Qiagen, Hilden, Germany) according to manufacturer’s instructions. Muscle gene expression was profiled with quantitative real-time RT–PCR using SYBR Green with validated primers. The relative expression of the selected genes was normalized to the reference gene TATA-Box Binde-Protein (TBP). A full list of primers is given in table S3.

**RNA Seq**

Total RNA was isolated using RNeasy Kit (Qiagen, Hilden, Germany) according to manufacturer’s instructions. For library preparation, 1 μg of total RNA per sample was used. RNA molecules were poly(A) selected, fragmented, and reverse transcribed with the Elute, Prime, Fragment Mix (EPF, Illumina). End repair, A-tailing, adaptor ligation, and library enrichment were performed as described in the TruSeq Stranded mRNA Sample Preparation Guide (Illumina). RNA libraries were assessed for quality and quantity with the Agilent 2100 BioAnalyzer and the Quant-iT Pico- Green dsDNA Assay Kit (Life Technologies). IMAT strand-specific RNA libraries were sequenced as 100+100 bp paired-end runs on an Illumina HiSeq2500 platform and muscle strand-specific RNA libraries were sequenced as 100+100 bp paired-end runs on an Illumina HiSeq4000 platform. The STAR aligner* (v 2.4.2a) with modified parameter settings (–twopassMode = Basic) was used for split-read alignment against the human genome assembly hg19 (GRCh37) and UCSC known Gene annotation. To quantify the number of reads mapping to annotated genes we used HTseq-count° (v0.6.1). RNA-Seq count files filtered for genes with an average read count above 50 counts per sample. DESeq2 [1] was used for expression level normalization using variance stabilizing transformation.

**Selection of Multivariate Regression hits**

Selection of gene specific regression models for predicting FG and GIR was done based on a combination of model log-likelihood and negative log-likelihood. Log likelihood was estimated with the *mvregress* Matlab function. Additionally, we calculated the negative log-likelihood for multivariate regressions using Matlab’s *mvregresslike* function for each model. Log-likelihood and negative log-likelihood distributions did not reveal a distinct cutoff criteria for model selection (Figure S1C). We thus manually applied a linear cutoff function to select a meaningful number of gene related models with high evaluation scores for both likelihood distributions (Figure S1C).

[1] Love, M.I., W. Huber, and S. Anders, *Moderated estimation of fold change and dispersion for RNA-seq data with DESeq2.* Genome Biol, 2014. **15**(12): p. 550.
